# Does objective feedback decrease sedentary behavior in geriatric rehabilitation? A systematic review

**DOI:** 10.1101/2025.06.24.25327890

**Authors:** Suzanne M. Debeij, Miriam L. Haaksma, Peter A. van de Hoef, Wilco P. Achterberg, Eléonore F. van Dam van Isselt

**Affiliations:** Department of Public Health and Primary care, Leiden University Medical Center, Leiden, the Netherlands; University Network for the Care Sector South-Holland, Leiden University Medical Center, Leiden, the Netherlands; LUMC Center for Geriatric Medicine, Leiden University Medical Center, Leiden, the Netherlands; Research Group Innovation of Human Movement Care, Research Center Healthy and Sustainable Living, HU University of Applied Sciences, Utrecht, the Netherlands; National eHealth Living Lab, Leiden, the Netherlands

**Keywords:** physical activity, post-acute care, older people, eHealth, remote (patient) monitoring

## Abstract

**Objective:** To assess the effectiveness of interventions using feedback from objective measurements to reduce sedentary behavior in older adults during rehabilitation.

**Data Source:** Cochrane Library, Web of Science, PubMed, Embase and Emcare were systematically searched for controlled trials in May 2023.

**Study Inclusion and Exclusion Criteria:** Studies in post-acute settings involving older individuals, providing feedback as part of the intervention and reporting effects on sedentary behavior were included.

**Data Extraction:** Study characteristics, aim, methodology, intervention content, and outcome measures with effect sizes relating to sedentary behavior were extracted.

**Data synthesis:** A narrative synthesis was used to describe the data.

**Results:** Eleven studies with diverse methods and reported outcomes were included. Feedback was focused only on sedentary behavior (one study), on sedentary behavior and physical activity (four studies), or only on physical activity (six studies). All studies showed high bias risk in at least one quality assessment domain. Six studies reported a significant decrease in sedentary behavior, while five reported no effect. No harms were reported.

**Conclusion:** The effect of feedback-based interventions on sedentary behavior in geriatric rehabilitation remains inconclusive. Interventions differed considerably in content and methodology, and high-quality evidence is lacking. Future research should focus on improving research methodology, optimizing strategies to reduce sedentary behavior and finding the minimum reduction of sedentary behavior required to create a clinically relevant difference.

**Key points:**

Although some studies indicate a (small) reduction in sedentary behaviour following interventions with objective feedback for older individuals in rehabilitation, effects remain inconclusive.
Interventions to decrease sedentary behaviour differ considerably in content and methodology.
Despite substantial evidence on the health risks associated with sedentary behaviour, there is a lack of high-quality studies on feedback to reduce sedentary time during rehabilitation.

## Introduction

Older individuals with disabling impairments due to stroke, exacerbation of COPD, a hip fracture, or other causes can be admitted to a rehabilitation department to improve their health. This type of post-acute care is an effective, patient-centered multidisciplinary approach, in which evaluative, diagnostic, and therapeutic interventions are integrated in a treatment plan^1, 2^. The tailored rehabilitation treatment plan aims to restore functional abilities, improve functional capacity, enhance social engagement, and elevate overall quality of life^1, 3^.

The older individuals in rehabilitation are encouraged to engage in physical activity to regain functional performance. However, they often do not have the capability to engage in physical activity by themselves or are unable to adhere to the activities^4^. Hence, patients in rehabilitation spend much of their time being sedentary^5, 6^. High sedentary time can have hazardous health effects such as increased risk of cardiometabolic conditions, and premature mortality^7^. Sedentary behavior can be defined as “any waking behavior characterized by an energy expenditure ≤1.5 Metabolic Equivalent of Task while in a sitting, reclining or lying posture”^8^. To describe sedentary behavior, for example, the total amount of sitting time, duration of sedentary breaks and length of uninterrupted bouts spent sedentary are used^9^. Particularly the latter tends to be detrimental to health^10^. Sedentary behavior differs from physical inactivity, as someone who sits most of the day, can still meet the WHO guidelines on the recommended amount of physical activity^7, 11^. Nonetheless, to counteract the risks of a high sedentary time, exponentially more time should be spent on physical activity, which is not feasible for older individuals with impairments^12^. Decreasing or changing the pattern of sedentary time may be a more feasible goal for these individuals to start health behavior change^7, 13^.

There are numerous models and techniques to guide or describe behavior change. Michie et al. (2013) created a cluster-divided taxonomy for behavior change techniques^14^. These clusters can be incorporated in existing behavior change models such as the behavior change wheel^14, 15^. One of the clusters of behavior change techniques is ‘feedback and monitoring’ and comprises techniques including feedback on behavior and self-monitoring of behavior^14^. Aspects from this and other clusters are known to reduce sedentary behavior in adults and older adults^16–19^.

Feedback and self-monitoring on objectively measured sedentary behavior can be provided and supported by eHealth, for example, on websites, in applications, and via wearables such as smartwatches and smartphones. In addition, a wearable or other forms of eHealth can be used independently of a healthcare provider which may increase access to services, save time and reduce costs^20^. Wearables could sense movement in the older people in rehabilitation objectively and unobtrusively via accelerometers. The accelerometers give more reliable data on sedentary time and other movement behaviors than self-reported data, which often underestimate the true value^21^. The obtained data can be transformed immediately into visual graphics, which can give the caregiver and patient insight into the actual behavior. The advantages of eHealth provide interesting possibilities to reduce sedentary behavior. Both in a community-dwelling setting and a hospital setting, eHealth interventions are shown to decrease sedentary time in older adults when compared with control conditions^17, 22^. However, in a geriatric rehabilitation population, the effect of eHealth interventions with feedback to decrease sedentary behavior is still unclear. This systematic review aims to assess the effectiveness of interventions using feedback based on objective measurements to decrease sedentary behavior of older adults in geriatric rehabilitation.

## Methods

### Data sources

For this systematic review, Cochrane Library, Web of Science, PubMed, Embase and Emcare were searched on May 30 and May 31 of 2023. The systematic review is reported based on the PRISMA 2020 guidelines and is pre-registered in PROSPERO (protocol number: CRD42023428935)^23, 24^.

### Inclusion and Exclusion Criteria

To be included in this review, studies had to be a randomized controlled trial or have a (quasi)experimental design. Studies were included if providing feedback from an objective measurement was part of the intervention, the intervention started within three months after an acute-care event and had objectively measured sedentary behavior as operationalized in the CROSBI consensus study (sedentary time, breaks and bouts and the number of transitions) as primary or secondary outcome^9^. Furthermore, studies had to include older individuals (mean age 55 years or older). Studies were not excluded based on study quality. The complete search string can be found in supplemental material 1.

### Data extraction and synthesis

All titles and abstracts identified by the search were uploaded in the systematic review management tool Covidence and duplicates were excluded. The title and abstracts of the potentially relevant studies were independently screened by two researchers (SD and MH). Next, the same researchers obtained and reviewed full texts to finalize inclusion. Disagreements between researchers were discussed until consensus was reached. If consensus could not be reached, a third researcher (LD) was consulted.

The first part of the data extraction (7 studies (64%)) was conducted independently by two researchers (SD and MH). The remaining data extraction (4 studies (36%)) was carried out by one researcher (SD) who discussed uncertainties with another researcher (MH). Full data extraction can be seen in supplemental materials 2 and 3. Participants’ demographics, study aim, design, method, intervention content, and outcome measures and effect sizes relating to sedentary behavior were described in a narrative synthesis.

### Quality assessment

The quality of the outcomes in the included randomized studies was assessed per outcome using the Cochrane risk of bias tool for randomized controlled trials (RoB2)^25^. The quality of outcomes in the non-randomized studies was assessed using The Risk Of Bias In Non-randomized Studies – of Interventions (ROBINS-I) assessment tool^26^.

## Results

### Study selection

The search identified 6231 studies. The PRISMA flowchart in figure 1 shows the results obtained at various stages. After exclusion based on title and abstract, 52 studies remained. With full text screening, another 41 studies were excluded. Finally, 11 studies were included in this review. A third researcher was needed to reach consensus in two cases. In one other case, the author of a study was contacted to clarify eligibility of the study.

**Figure 1.**
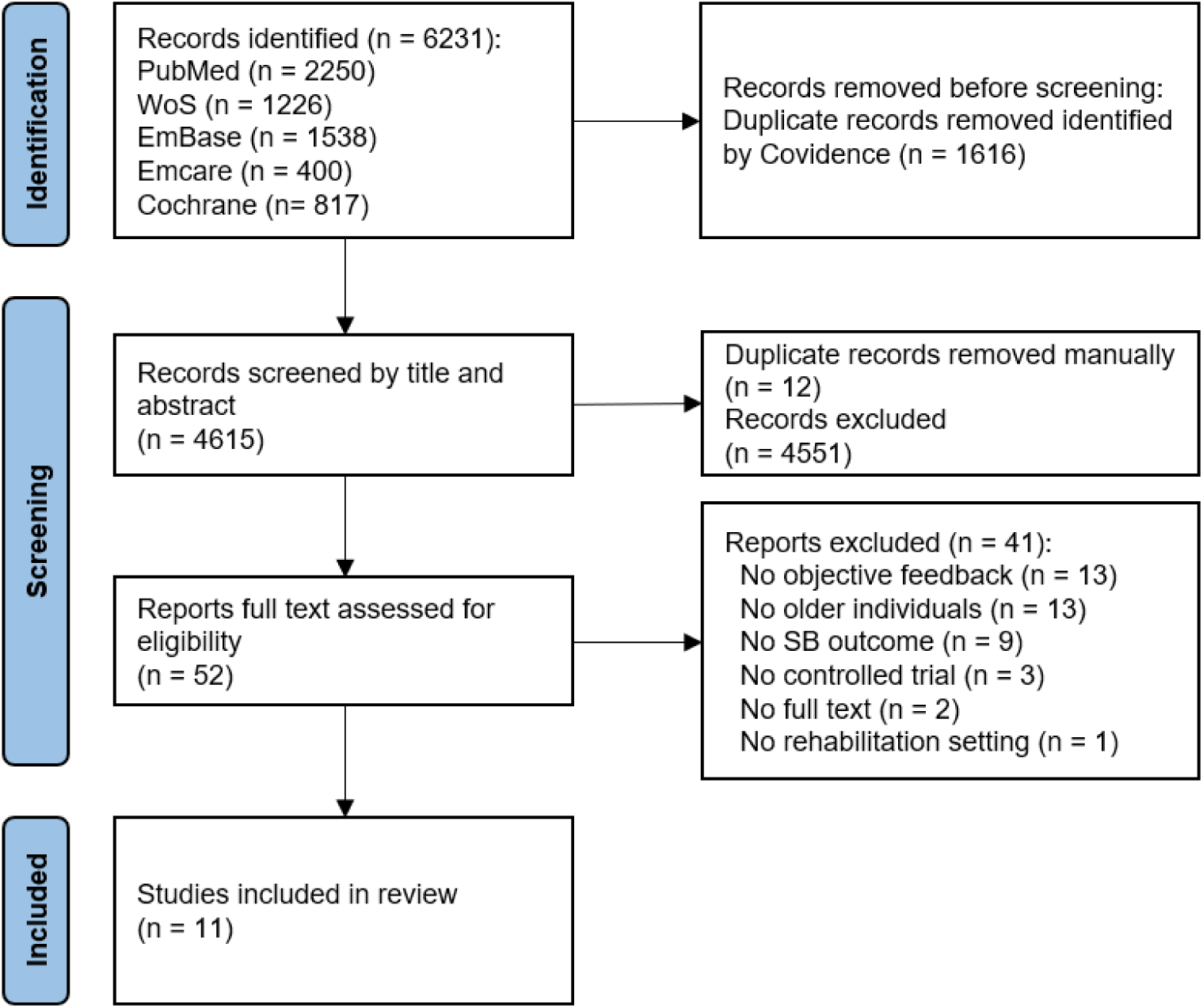
PRISMA flow diagram of search strategy results.

### Study characteristics

Nine of the eleven (82%) included studies were randomized controlled trials 27-35 and two (18%) were historically controlled trials 36,37. As shown in table 1, four of the included studies were conducted in a rehabilitation setting 29,30,32,34, four in a hospital setting 31,35-37, two during hospitalization and after discharge 27,28, and one in a community setting 33. Two of the included articles were based on the same intervention study and had partial overlap in participants 27,28. The difference between these articles was the follow-up time of the intervention.

**Table 1.** Characteristics of included studies.

| Author (year) | Country | Design | Setting | Diagnosis | Study duration | Sample size | mean age | %female |
| --- | --- | --- | --- | --- | --- | --- | --- | --- |
| Ashizawa et al. (2022)<sup>29</sup> | Japan | RCT | Hospital and post-discharge | Stroke | hospitalization + 3 months | 61 | 71 | 34 |
| Ashizawa et al. (2023)<sup>28</sup> | Japan | RCT | Hospital and post-discharge | Stroke | hospitalization + 6 months | 86 | 72 | 33 |
| Atkins, Canell and Barr (2019)<sup>32</sup> | Australia | RCT | Subacute hospital rehabilitation unit | Mobility | ± 1 month (admission - discharge) | 78 | 76 | 59 |
| Brandes et al. (2018)<sup>33</sup> | Germany | RCT | Inpatient rehabilitation | Joint | ± 3 weeks + 6 months | 49 | 70 | 53 |
| Conijn et al. (2020)<sup>34</sup> | the Netherlands | HCT | Hospital | Diverse | hospitalization + 1 month | 94 | 58 | 44 |
| Dall et al. (2019)<sup>30</sup> | Denmark | RCT | Hospital | Pulmonary | 1 - 7 days | 93 | 73 | 51 |
| Hassett et al. (2020)<sup>31</sup> | Australia | RCT | Neurological inpatient rehabilitation | Diverse | 6 months | 300 | 72 | 50 |
| Ozemek et al. (2020)<sup>35</sup> | United States | RCT | Phase III maintenance CR program | Cardiac | 3 months | 74 | 61 | 23 |
| Ter Hoeve et al. (2018)<sup>36</sup> | the Netherlands | RCT | Outpatient cardiac rehabilitation center | Cardiac | 3 months (+ 9 months aftercare) | 324 | 59 | 20 |
| van Bakel et al. (2023)<sup>27</sup> | the Netherlands | RCT | Hospital | Cardiac | 3 months | 212 | 63 | 23 |
| van Dijk - Huisman et al. (2020)<sup>37</sup> | the Netherlands | HCT | Hospital | Joint | median 3 days | 97 | 66 | 43 |

*RCT = randomized controlled trial, HCT = historically controlled trial*

Three of the studies included participants with a cardiac-related diagnosis 33-35, two with stroke 27,28, two with joint-related diseases 30,37, two with a diverse participant group 32,36, one with participants with mobility issues 29 and one with pulmonary diseases 31. The study duration varied from under one week 31 to over six months 28,29,34. The sample size across the studies ranged from 49 to 324 participants. The mean age ranged from 58 years 36 to 76 years 29. All study characteristics are reported in table 1.

### Interventions

One study described how the intervention was designed together with patients and provided feedback only on sedentary behavior (table 2)^27^. The other studies simply described the intervention, not how it was constructed. Four studies described interventions that provided feedback on sedentary behavior and physical activity^28–31^ and six provided feedback only on physical activity^32–37^ (table 1). Feedback frequency varied from continuous feedback available throughout the day to feedback twice a week. The duration of the intervention period varied as well, from a few days only in a clinical setting^30, 37^ to up to a full year both in the clinical setting and after discharge^36^. The form in which feedback was provided also varied between studies. Ten studies described feedback being given by a digital device^27–32, 34–37^. Six of these studies also incorporated feedback from a professional in the intervention^27–29, 31, 32, 37^. The feedback from professionals was provided via phone calls in four studies^27–29, 31^ in-person in two studies^32, 37^ and via both routes in two studies^28, 29^. In one study, feedback was not provided by a digital device directly, but only via graphic visualization during the physical activity counselling sessions^33^. Furthermore, the feedback was discussed or evaluated with caregiver(s) in eight studies^27–29, 31, 33, 34, 36, 37^. The other three studies did not set-up the intervention as blended-care but used feedback as an addition to the care-pathway^30, 32, 35^.

**Table 2.** Intervention and measurements from included studies.

| Author (year) | Description of intervention | Control group treatment | Feedback |  |  |  | Sensor for outcome measurement |
| --- | --- | --- | --- | --- | --- | --- | --- |
|  |  |  | *Duration* | *Provided via* | *Frequency* | *Given on* |  |
| Ashizawa et al. (2022)<sup>28</sup> | During hospitalization: educational pamphlet on reducing SB, goal setting for SB after discharge and self-monitoring of SB and step count.<br>After discharge: self-monitoring of SB and step count, motivational stickers and phone calls for encouragement and feedback | During hospitalization: educational pamphlet on increasing PA, self-monitoring of step count. Feedback on checklist once a week<br>After discharge: self-monitoring of steps | hospitalization + 3 months | Active Style Pro HJA-750, and by professional (by phone or face-to-face) | Daily by self-monitoring,<br>Once a week from professional during hospitalization, once every two weeks after hospitalization | SB, PA, and step count | Active Style Pro HJA-750C |
| Ashizawa et al. (2023)<sup>29</sup> | During hospitalization: educational pamphlet on reducing SB, goal setting for SB after hospital discharge and self-monitoring of screen time and step count<br>After discharge: self-monitoring of screen time and step count, motivational stickers and phone calls for encouragement and feedback | During hospitalization: educational pamphlet on increasing PA, self-monitoring of step count. Feedback on checklist once a week<br>After discharge: self-monitoring of steps | hospitalization + 3 months | Active Style Pro HJA-750C, and by researcher (by phone or face-to-face) | Daily by self-monitoring,<br>Once a week from professional during hospitalization, once every two weeks after hospitalization | SB and step count | Active Style Pro HJA-750C |
| Atkins, Canell and Barr (2019)<sup>32</sup> | An exercise program, feedback from the pedometer and small phrases of encouragement | Usual care and an exercise program, estimation of distance walked each day | ± 1 month (admission - discharge) | Yamax Digiwalker SW200 and by family and nursing staff | Continuous feedback and small phrases of encouragement | Step count | ActivPAL Yamax Digiwalker SW200 |
| Brandes et al. (2018)<sup>33</sup> | Encouragement to increase daily step count by 5%, PA counselling twice a week to encourage patients to use restored walking capabilities | Standard rehabilitation | ± 3 weeks (inpatient rehabilitation) | Graphical visualizations of steps taken shown by PA counsellors | Twice a week in a counselling session | Step count | Step Activity Monitor 3.0 (SAM 3.0) |
| Conijn et al. (2020) <sup>34</sup> | A multicomponent intervention which included paper and digital information about the importance of PA, an exercise movie, an activity planner, a pedometer and a wearable activity tracker, personal activity coaching and access to a tailored digital exercise program | Usual care | hospitalization + 1 month | Fitbit Flex and pedometer | Continuous during hospitalization and 1 week after discharge | PA and step count | Activ8 |
| Dall et al. (2019) <sup>30</sup> | Feedback on time spent lying in bed, sitting, standing, and walking via an app on a tablet on bedside table. The app contained 'smiley-faces' in different colors. Feedback was visible for patients, visitors, and healthcare personnel. | Usual care | 1 - 7 days | An app connected to 2 small tri-axial accelerometers embedded in medical Band-Aids | Continuous | Time bedridden, sitting, standing, and walking | 2 small tri-axial accelerometers embedded in medical Band-Aids |
| Hassett et al. (2020) <sup>31</sup> | Tailored digitally-enabled rehabilitation with exercises, remote monitoring and communication, and a fall prevention brochure | Usual care, prescription of repetitive exercises, tailored management, and a fall prevention brochure | 6 months | Activity monitor (Fitbit, Garmin), apps and from therapist (by phone) | Continuous and by phone as required | Performance (dependent on device used; PA, SB or other) | ActivPAL |
| Ozemek et al. (2020) <sup>35</sup> | PF group: weekly goal to increase PA by 10%, if goal was not met the goal was maintained. Weekly booklets to increase PA. PF+MM group: additional to PF intervention receiving a newsletter on their (not) accomplished PA goal. | Usual care, recommendations to perform home exercises on ≥2 non-CR days | 3 months | Liferecorder PLUS, and via newsletter | Continuous, write down 4 times a day | PA goal (step count) | Actigraph GT3X |
| Ter Hoeve et al. (2018) <sup>36</sup> | 3 group PA counselling sessions, pedometers for daily PA feedback and goal setting with physical therapist, booklet with assignments, information on goal setting, barrier identification and relapse prevention, information about breaking up SB time. | Usual care, educational sessions, group counselling if indicated, information about general health benefits of PA, no aftercare | 3 months (+ 9 months aftercare) | Yamax Digiwalker SW200, self-monitoring | Continuous during CR, 3 times during after-care | PA (step count), aerobic capacity | Actigraph GT3X |
|  | In aftercare 3 sessions of PA and counselling |  |  |  |  |  |  |
| van Bakel et al. (2023) <sup>27</sup> | Patient education, goal setting, motivational interviewing with coping planning, (tele)monitoring | Usual care | 3 months | Web-based dashboard, smartphone application, vibrotactile feedback from Activ8 and professional (by phone) | Continuous, and (bi)weekly telephone coaching | SB and ST | ActivPAL3 |
| van Dijk - Huisman et al. (2020) <sup>37</sup> | Feedback for patient and physiotherapist on time spent standing and walking, feedback on functional recovery assessment, personalized exercise program | Usual care | Median 3 days | Smartphone application coupled with MOX activity monitor and by professional | Continuous | Time spent standing/walking | MOX activity monitor |

*SB = sedentary behavior, PA = physical activity, PF = pedometer feedback, PF + MM = pedometer feedback + motivational messaging, IPAQ = International physical activity questionnaire, CR = cardiac rehabilitation*

Nine different types of sensors were used to objectively measure sedentary behavior, as described in table 2. The ActivPAL^31, 32^, ActiGraph GT3X^35, 36^ and Active Style Pro HJA 570-C^28, 29^ were used in two studies. Other sensors - Step Activity Monitor 3.0, Activ8, accelerometers in medical Band-Aids, ActivPAL3 and MOX activity monitor - were used in one study.

### Study quality

The results of the quality assessments are presented in figure 2. Because multiple outcomes within one study scored the same risk level in all domains of the quality assessment, only one assessment per study is shown. Although four RCTs scored low risk in four out of five domains^27, 30, 31, 36^, all included studies scored an overall high risk of bias. The most frequent shortcoming was seen in the domain *missing outcome data*, where only one study scored a low risk of bias^27^. By contrast, the domain *measurement of the outcome* scored low risk in all studies. In the domain *deviations from the intended interventions* two studies scored a high risk^27, 32^. Atkins et al. (2019)^32^ reported a contamination of study results, because some participants in the intervention group were aware of their step count. Van Bakel et al. (2023)^27^ also reported on the risk of contamination between intervention and control group. Furthermore, three studies scored low risk in three domains^28, 29, 32^ and only one study scored a high risk in more than one of the domains^33^.

**Figure 2.**
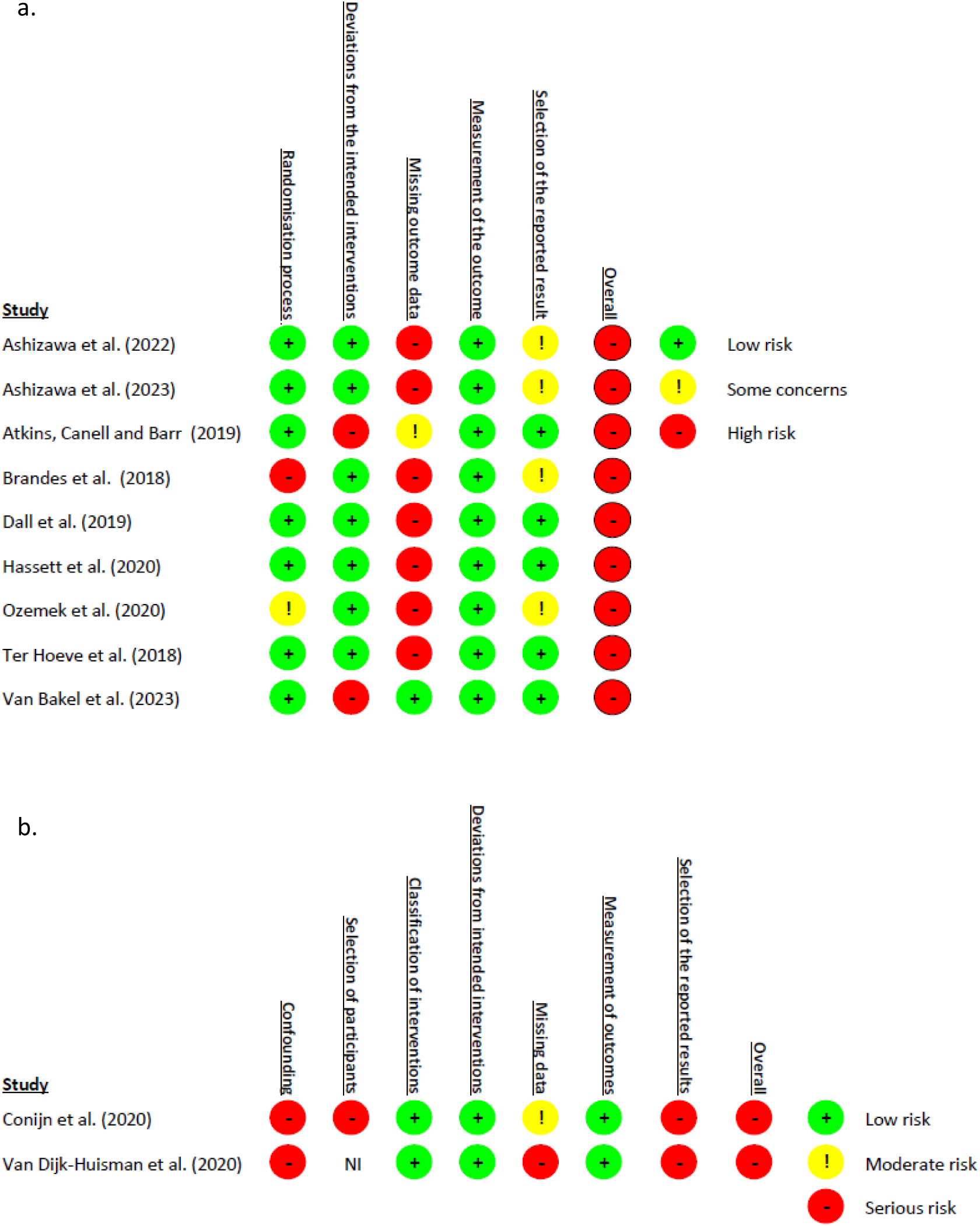
Quality assessments of the studies. a) RoB2 quality assessments, b) Robins-I quality Assessments *NI = No Information*.

The overall score of the quality assessment of both historically controlled trials was serious^34, 37^. Both studies scored a serious risk in the domains of *confounding* and *selection of the reported results*. Further, both did not control for all the important confounding domains (for example multimorbidity). In the domain of *bias due to selection of the reported results*, it was likely that reported estimates in both studies were selected from multiple analyses.

### Effect of feedback on sedentary behavior

Studies operationalized sedentary behavior outcomes differently. The most frequently reported outcome was the change in sedentary behavior/time (55%) ^27–29, 34–36^. Details can be found in table 3. As two RCTs failed to report a between-group comparison ^33, 35^, a distinction has been made between studies that reported differences between intervention and control groups, and studies that only reported differences within intervention and control groups.

**Table 3.** Sedentary outcomes.

| Feedback on | SB outcome measure | Unit | Author (year) | Estimate (95% CI)<br>*Intervention minus control* | p value | Study conclusion |
| --- | --- | --- | --- | --- | --- | --- |
| Outcomes compared between intervention and control |  |  |  |  |  |  |
| **SB** | Sedentary time | Hours/day | van Bakel et al. (2023)<sup>27</sup> | -0.4 h (-1.0, 0.3) | P=0.27 | Although the intervention was feasible, there was no evidence of effectiveness of the intervention |
|  | Number of prolonged sedentary bouts | Number/day | van Bakel et al. (2023)<sup>27</sup> | -0.1 (-0.8, 0.5) | p=0.71 | The number of prolonged sedentary bouts reduced, but change was comparable between groups. |
| **SB & PA** | Sedentary time | Percentage change | Ashizawa et al. (2022)<sup>29</sup> | -7.9%, | p=0.018 | An approach that encourages SB reduction reduces SB more than an approach that promotes PA. |
|  |  |  | Ashizawa et al. (2023)<sup>28</sup> | PI: -7.8% (-13.88, -1.65)<br>FU: -6.8% (-12.67, -0.88) | PI: p=0.013<br>FU: p=0.025 | Encouragement of reducing SB is effective up to 6 months after discharge |
|  | Sitting time | Minutes/day | Dall et al. (2019)<sup>30</sup> * | 52 min (-118, 23) | p=0.54 | There was no evidence of effectiveness of the intervention. |
|  | Time spent in bed | Minutes/day | Dall et al. (2019)<sup>30</sup> * | -70 min (-265, 125) | p=0.48 | There was no evidence of effectiveness of the intervention. |
|  | Sit-to-stand transitions | Number/day | Hassett et al. (2020)<sup>31</sup> | 3 weeks: 0 (-4, 3)<br>6 weeks: 2 (-2, 6) | 3 weeks: p=0.88<br>6 weeks: p=0.31 | There was no evidence of effectiveness of the intervention. |
| **PA** | Sedentary time | Interaction effect | Conijn et al. (2020)<sup>34</sup> * | DH: (-1.57, 0.17) +<br>AD: (-1.36, 1.00) | DH: p=0.11<br>AD: p=0.76 | IG tend to (not significant) spend less time sedentary in hospital, not after discharge. |
|  |  | Percentage change | Ter Hoeve et al. (2018)<sup>36</sup> | B=0.39 (0.82, 1.59) | p=0.53 | There was no evidence of effectiveness of the intervention. |
|  | Sedentary bouts ≥ 30 min | Percentage change | Ter Hoeve et al. (2018)<sup>36</sup> | B=0.76 (1.02, 2.53) | p=0.40 | There was no evidence of effectiveness of the intervention. |
|  | Time spent upright | Hours | Atkins, Canell and Barr (2019)<sup>32</sup> * | 0.55 hrs. | p=0.004 | Reading steps from pedometers increases the time spent upright in first week of rehabilitation. |
|  | PA on POD1 | Minutes | van Dijk - Huisman et al. (2020)<sup>37</sup> * | B 28.43 (5.55, 51.32)<br>SE 11.50 | p=0.016 | The intervention shows potential to enhance PA levels and recovery process |
| Outcomes compared pre-post within a group |  |  |  |  |  |  |
| **PA** | Sedentary time | Percentage change | Ozemek et al. (2020)<sup>35</sup> | CG: 1.3%<br>PF: 2.6%<br>PF+MM: 1.8% | CG: p=0.29<br>PF: P= 0.03<br>PF+MM: p=0.19 | CR programs are encouraged to implement PA feedback with individualized PA goals to support the increase in PA and potentially decrease SB. |
|  |  | Minutes/day |  | C: 14.6 min<br>PF: 22.4 min<br>PF + MM: 6.5 min | UC: p=0.34<br>PF: p=0.01<br>PF+MM: p=0.54 |  |
|  | Inactive time | Minutes | Brandes et al. (2018)<sup>33</sup> | Onset-1m<br>IG: -10 min<br>CG: -9.5 min<br>Onset-6m<br>IG: -54 min<br>CG: -29.3 min | onset-1m<br>IG: p>0.05<br>CG: p>0.05<br>onset-6m<br>IG: p<0.05<br>CG: p>0.05 | The intervention group significantly decreased the inactive time 6 months post rehabilitation compared to one month post rehabilitation. |
|  | Mean duration of inactive bouts | Minutes | Brandes et al. (2018)<sup>33</sup> | Onset-1m<br>IG: -1.2 min<br>CG: 0 min<br>Onset-6m<br>IG: -2 min<br>CG: -2.6 min | onset-1m<br>IG: p>0.05<br>CG: p>0.05<br>onset-6m<br>IG: p>0.05<br>CG: p>0.05 | IG and CG both did not significantly reduce the mean duration of inactive bouts. |
|  | Maximum duration of inactive bouts | Minutes | Brandes et al. (2018)<sup>33</sup> | Onset-1m<br>IG: -17.4 min<br>CG: -7.2 min<br>onset-6m<br>IG: -28.4<br>CG: -30.8 | onset-1m<br>IG: p>0.05<br>CG: p>0.05<br>onset-6m<br>IG: p>0.05<br>CG: p>0.05 | IG and CG both did not significantly reduce the maximum duration of inactive bouts. |

\*Study did not report on change over time, † study did not report effect estimate, only the 95% CI, SB = sedentary behavior, PA = physical activity, PI = post intervention, FU = follow-up, PA = physical activity, DH = during hospitalization, AD = after discharge SE = standard error, IG = intervention group, CG = control group, PF = pedometer feedback group, PF+MM = pedometer feedback + motivational messaging group, CR = cardiac rehabilitation, POD1 = post operative day 1, 1m = 1 month, 6m = 6 months

### Differences between intervention and control

Four of the nine studies that compared outcomes between intervention and control group reported a significant effect on sedentary behavior ^28, 29, 32, 37^. As shown in table 3, only in the intervention of van Bakel et al. (2023) feedback was focused solely on sedentary time ^27^. The authors reported a decrease of 0.4 hours/day more in the intervention group than in the control group, which was non-significant. However, the intervention was described as feasible and promising because it reduced the odds of sitting longer than 9.5 hours/day. Two of the four studies that provided feedback on both physical activity and sedentary behavior reported even more positive results; both reported a reduction of almost 8% of sedentary time in the intervention group post intervention ^28, 29^. These two studies from Ashizawa et al. concluded that an approach that encourages sedentary behavior reductions is more effective than an approach that promotes an increase in physical activity for patients with minor ischemic stroke. However, the third study with an intervention that provided feedback both on physical activity and sedentary behavior, the study of Dall et al. (2019), reported an increased sitting time of 52 minutes/day in the intervention group ^30^. Nevertheless, the total time spent in bed was 70 minutes/day less in the intervention group. Since both differences were not significant, the study did not report the intervention as effective. The fourth study reported a non-significant mean reduction of less than three sit-to-stand transitions in the intervention group compared to the control group ^31^.

The other four studies describe interventions in which participants received feedback on various forms of physical activity ^32, 34, 36, 37^. Ter Hoeve et al. (2018) concluded that the intervention with counselling sessions was not successful in changing sedentary behavior, as the intervention group and control group barely showed a difference in sedentary time (0.39% (-.82, 1.59)) ^36^. Conijn et al. (2020) did not report on change over time and described that the intervention group tended to spend less time sedentary during hospitalization, but not in the first week after discharge while they still followed the intervention ^34^. They reported that their results should be interpreted with caution, as they struggled with methodological issues such as a high drop-out rate. The remaining two studies also did not report on change over time but did report a significant effect between the control group and intervention group on the sedentary behavior outcome in the measured period ^32, 37^. The median time spent upright was 0.55 hours higher than the median of the control group ^32^. Van Dijk-Huisman et al. (2020) ^37^ reported that the intervention group was 28.4 minutes more active than the control group.

### Outcomes compared pre-post within a group

Two studies reported pre-post results within groups ^33, 35^ instead of between-groups comparisons. Both interventions contained feedback on PA. The study of Brandes et al. (2018) ^33^ reported a statistically significant decrease of 54 inactive minutes from onset to six months post-rehabilitation in the intervention group. The decrease of 29.3 minutes in the control group over the same time period was not significant. The mean duration of inactive bouts and maximum duration decrease in both groups. The study of Ozemek et al. (2020) ^35^ did report a significant decrease in sedentary time of 2.6%, or 22.4 (12.8) minutes in the intervention group that received feedback using a pedometer.

## Discussion

This review provides an overview of the effect of feedback from an objective measurement for older adults in geriatric rehabilitation on their sedentary behavior. Included studies varied greatly in both interventions (such as backgrounds of participants, the way feedback was provided and duration of the intervention) as in the follow-up, outcome measures and units that were reported. Besides, most studies were of poor quality. In six studies, the authors described a significant decrease in a sedentary behavior outcome^28, 29, 32, 33, 35, 37^. The other five studies did not report a statistically significant difference^27, 30, 31, 34, 36^. Two of the eleven studies did not examine the preferred contrast of interest by conducting only within-group comparisons^33, 35^. None of the studies reported a harmful or negative effect of the intervention. Evidence for the effectiveness of an intervention providing feedback based on objective measurement to decrease sedentary behavior remains inconclusive.

The methods used in studies that described an effect on sedentary behavior varied as much as studies that did not report a significant effect. Not one component appeared to be the most successful. A suggestion in the literature to increase effectiveness of an intervention to change behavior is the incorporation of more behavior change techniques or the incorporation of feedback into a multi-component intervention^38^. However, the number of intervention components in this review varied but interventions with more components did not show higher effectiveness than studies with fewer components. Perhaps it is not the number of the components of interventions that is important, but rather their tailored use^39^. With over 90 possible behavior change techniques, the person delivering the intervention should be aware of the personal preferences of a patient, which can depend on a person’s capabilities, the opportunities that are offered and the motivation to change^14, 15^. Studies in this review did not describe whether the interventions had taken these factors into account. The possible lack of a personalized approach may also explain why the frequency of the feedback and the form in which feedback was provided showed no effect on the outcome. Besides, the effectiveness of studies that incorporated the feedback as blended care, did not differ from the effectiveness of studies that provided feedback in addition to the care-pathway. This contrasts with the literature, which states that digital interventions in rehabilitation are more effective when provided as blended care^40^. In addition, the variation in outcomes could be explained by the difference in accelerometers that were used in the interventions. All accelerometers measure sedentary behavior slightly differently, and the validity of accelerometers is known to vary, especially for individuals with a divergent movement pattern (i.e., smaller step length or low walking speed)^41, 42^.

Studies in this review did not describe a harmful effect but were inconclusive regarding the effectiveness of feedback on reducing sedentary behavior in this target group. The studies that focused on sedentary behavior did not report a larger effect than studies with interventions focused on physical activity. This is in contrast to the literature, which suggests that interventions for adults with a focus on sedentary behavior instead of physical activity tend to have a more promising effect on decreasing sedentary behavior^43, 44^. The difference could be due to the geriatric rehabilitation target group in this study. Although patients had diverse backgrounds, they all received rehabilitation and were older individuals. However, despite the often-described barriers for older adults to use eHealth, such as low digital literacy, studies show that older patients can engage with eHealth during rehabilitation and that using eHealth can potentially improve rehabilitation outcomes^40, 45, 46^. Our inconclusive outcomes show similarities with those of other studies focusing on older adults. In community-dwelling older adults, interventions with a (digital) feedback component have the potential to reduce sedentary time, but did not achieve statistical significance^17^. According to the authors, the lack of statistical significance in this meta-analysis could be due to both the small number of included studies and the small sample size of existing studies. Studies in younger adults provide more evidence on reducing sedentary behavior by eHealth with a feedback component. For example, a scoping review reporting on the reduction of sedentary behavior with single component mHealth interventions in young and middle-aged adults reported a positive effect in seven of the nine studies^47^. A systematic review and meta-analysis on community-dwelling inactive young and middle-aged adults also provided evidence that mobile health interventions reduce sedentary behavior^48^. Overall, the literature tends to be mildly positive about the reduction of sedentary behavior but also states that much is unclear and that more high-quality studies are required.

Although all studies scored a high risk of bias on the quality assessment, the quality of the studies clearly varies. Four studies scored a high risk in only one domain, which resulted in an overall rated high-risk rating^27, 30, 31, 36^. However, they scored a low risk on the four other domains. These four studies all reported no effect of the intervention. Since all studies included in this review showed an overall high risk of bias, we can only speculate about the potential effect in a high-quality study. In some cases, the quality of included studies may even seem higher than it actually is, because clarity of results is not a domain in the quality assessment. The lack of this domain in some cases resulted in, for example, a low-risk score on the domain *selection of the reported results*, while limited information on the exact selection of results was reported. Based on the available data, derived solely from studies of suboptimal quality, higher quality studies did not show a larger effect compared to lower quality studies.

### Strengths and limitations

The extensive search strategy that covered five databases is the first strength of this systematic review. A second strength is the use of double-blind screening for study inclusion. A third strength is that this is the first review to report on the effectiveness of feedback on objectively measured sedentary behavior for older individuals in geriatric rehabilitation. Nonetheless, several limitations should also be noted. Part of the studies focused primarily on increasing physical activity rather than targeting sedentary behavior. This might have influenced their results on sedentary behavior. Second, the quantitative comparison across studies was hampered by the large variety in measurement methods and reported outcomes, complicating the synthesis of the results. Further, there was the heterogeneity in content, duration, and frequency of the administered interventions. Therefore, no meta-analyses could be conducted on this topic.

### Conclusions

Although some studies with older individuals in geriatric rehabilitation indicate a reduction in sedentary behavior following interventions with objective feedback, the effects remain undetermined. Clear evidence is lacking as only a limited number of studies could be included in this review and almost half of the studies did not find a positive effect on sedentary behavior. All included studies had a high risk of bias. Additionally, some studies did not report on the change over time or only conducted within-group comparisons. Remarkably, no indications regarding which type, duration or content of the interventions would be most optimal to affect the outcome regarding sedentary behavior could be found. To optimize rehabilitation for older people, high-quality randomized controlled trials are essential. Future research should focus on gaining insight into (1) the best method for providing feedback on sedentary behavior, and (2) the decrease in the total volume and bouts of sedentary behavior required to achieve clinical relevance.

## Data Availability

All data produced in the present work are contained in the manuscript.

## Acknowledgements

The authors are grateful to Pema Gurung for advising on the search string for this study and Maggie Oates for language editing. During the preparation of this work the authors used ChatGPT to improve their writing style. After using this tool, the authors reviewed and edited the content as needed and take full responsibility for the content of the publication.

## Declaration of conflicting interest

The authors declare no conflict of interest.

## Funding statement

This work was supported by the University Network for the Care Sector South-Holland.

## Ethical considerations

Not applicable

## Consent to participate

Not applicable

## Consent for publication

Not applicable

## Notes

### Competing Interest Statement

The authors have declared no competing interest.

### Clinical Protocols

https://www.crd.york.ac.uk/PROSPERO/view/CRD42023428935

