## Supplemental material 1 for "Does objective feedback decrease sedentary behavior in geriatric rehabilitation? A systematic review"

**PubMed**

*Search carried out on 30-05-2023*

| Search | Query | Results | Time |
|---|---|---|---|
| #5 | Search: **#1 AND #2 AND #3 AND #4** | 2,249 | 03:40:28 |
| #4 | Search: **"Physical and Rehabilitation Medicine"[Mesh] OR "Rehabilitation"[Mesh] OR rehab*[tiab] OR erehab*[tiab] OR "Physical Therapy Modalities"[Mesh] OR "physical therap*"[tiab] OR physiotherap*[tiab] OR "Rehabilitation Centers"[Mesh] OR "Subacute Care"[Mesh] OR subacute[tiab] OR postacute[tiab] OR "post-acute"[tiab] OR "Postoperative Period"[Mesh] OR "Aftercare"[Mesh] OR "Postoperative Care"[Mesh] OR "postoperative"[tiab] OR "post-operative"[tiab] OR inpatient[tiab] OR "sub-acute"[tiab] OR "hospital care"[tiab] OR aftercare[tiab] OR "followup care"[tiab] OR "follow-up care"[tiab]** | 1,384,871 | 03:40:13 |
| #3 | Search: **("Mobile Applications"[Mesh] OR "Appstore"[tiab] OR "App store"[tiab] OR "Google play*"[tiab] OR "Googleplay"[tiab] OR "Playstore"[tiab] OR "Play Store"[tiab] OR "ehealth"[tiab] OR "e-health"[tiab] OR "mhealth"[tiab] OR "m-health"[tiab] OR "e-rehab*"[tiab] OR "e-exercis*"[tiab] OR "personal mobility device*"[tiab] OR (("electronic"[tiab] OR "online" OR web[tiab] OR internet[tiab]) AND (platform*[tiab] OR wearable*[tiab])) OR "webbased"[tiab] OR "web based"[tiab] OR "digital software"[tiab] OR smartwatch*[tiab] OR (application*[tiab] OR App[tiab] OR apps[tiab]) AND (mobile[tiab] OR "smartphone"[Mesh] OR smartphone*[tiab] OR Apple[tiab] OR Samsung[tiab] OR Android[tiab])) OR "Accelerometry"[Mesh] OR "Accelerometry"[tiab] OR "Accelerometer*"[tiab] OR " Pedometer*"[tiab] OR " inclinometer*"[tiab] OR "Actigraphy"[tiab] OR "activity monitor"[tiab] OR "step counter"[tiab]** | 79,770 | 03:40:04 |
| #2 | Search: **"Sedentary Behavior"[Mesh] OR "Health Behavior"[Mesh] OR "Television"[Mesh] OR "sedentary behaviour*"[tiab] OR "Sedentary Behavior*"[tiab] OR "Sedentary Lifestyle"[tiab] OR "Physical Inactivity"[tiab] OR "Lack of Physical Activity"[tiab] OR "Sedentary Time"[tiab] OR "Sedentary Times"[tiab] OR "Sitting"[tiab] OR "Screen Time"[Mesh] OR "screen times"[tiab] OR "inactive"[tiab] OR "Sedentary"[tiab] OR "watching TV"[tiab] OR "TV watching"[tiab] OR "viewing TV"[tiab] OR "TV viewing"[tiab] OR "television watching"[tiab] OR "watching television"[tiab] OR "television viewing"[tiab] OR "viewing television"[tiab] OR "screen-based "[tiab] OR "media time"[tiab] OR "Physical activit*"[tiab] OR "Exercise"[Mesh] OR "Exercise"[tiab]** | 1,022,973 | 03:39:55 |

| Search | Query | Results | Time |
| --- | --- | --- | --- |
| #1 | Search: **("Aged"[majr] OR "Aged, 80 and over"[majr] OR "frail elderly"[majr] OR "elderly"[tw] OR "elder"[tw] OR "elders"[tw] OR "aged"[ti] OR "aging"[tw] OR "ageing"[tw] OR "oldest old"[tw] OR "older person*"[tw] OR "old person*"[tw] OR "older patient*"[tw] OR "old patient*"[tw] OR "older women"[tw] OR "old women"[tw] OR "older men"[tw] OR "old men"[tw] OR "old adult*"[tw] OR "older adult*"[tw] OR "Older individual*"[tw] OR "old people"[tw] OR "older people"[tw] OR "septuagenarian*"[tw] OR "octagenarian*"[tw] OR "octogenarian*"[tw] OR "nonagenarian*"[tw] OR "centenarian*"[tw] OR "senescence"[tw] OR "senescent"[tw] OR "geriatric"[tw] OR "geriatrics"[tw] OR "geriatrics"[majr] OR "senior"[tw] OR "seniors"[tw] OR "older population"[tw] OR "neurogeriatr*"[tw] OR "Sixty plus"[tw] OR "Middle Aged"[Mesh] OR "60 years old"[tw] OR "60 age"[tw] OR "63 years old"[tw] OR "63 ±"[tw])** | 5,804,287 | 03:39:34 |

**Embase**

Embase <1974 to 2023 May 30>

1 exp sedentary lifestyle/ 20314
2 exp health behavior/ 492423
3 *television viewing/ 978
4 *screen time/ 1090
5 *exercise/ 130662
6 *physical activity/ 53497
7 ("sedentary behavio?r*" or "Sedentary Lifestyle" or "Physical Inactivit*" or "Lack of Physical Activit*" or "Sedentary Time*" or "Sitting" or "screen time*" or "inactive" or "Sedentary" or "watching TV" or "TV watching" or "viewing TV" or "TV viewing" or "television watching" or "watching television" or "television viewing" or "viewing television" or "screen-based " or "media time" or "Physical activit*" or "Exercise").ti,ab,kw. 759826
8 1 or 2 or 3 or 4 or 5 or 6 or 7 1237525
9 *geriatrics/ 23302
10 exp aged/ 3594777
11 exp very elderly/ 285696
12 ("Aged" or "Aged, 80 and over" or "frail elderly" or "elder*" or "aging" or "ageing" or "oldest old" or "older person*" or "old person*" or "older patient*" or "old patient*" or "older women" or "old women" or "older men" or "old men" or "old adult*" or "older adult*" or "Older individual*" or "old people" or "older people" or "septuagenarian*" or "octagenarian*" or "octogenarian*" or "nonagenarian*" or "centenarian*" or "senescence" or "senescent" or "geriatric" or "geriatrics" or "geriatrics" or "senior" or "seniors" or "older population" or "neurogeriatr*" or "Sixty plus" or "Middle Aged" or "60 years old" or "60 age" or "63 years old").ti,ab,kw.1984586
13 9 or 10 or 11 or 12 4809674
14 mobile application/ or exp telehealth/ or exp telerehabilitation/ or *accelerometer/ or *accelerometry/ or *pedometer/ or *actimetry/ or *activity tracker/ 112176
15 ("Appstore" or "App store" or "Google play*" or "Googleplay" or "Playstore" or "Play Store" or "ehealth" or "e-health" or "mhealth" or "m-health" or "e-rehab*" or "e-exercis*" or "personal mobility device*" or (("electronic" or "online" or web or internet) adj (platform* or wearable*)) or "webbased" or "web based" or "digital software" or smartwatch* or ((application* or App or apps) adj (mobile or "smartphone" or smartphone* or Apple or Samsung or Android))).ti,ab,kw. 87383
16 ("Accelerometry" or "Accelerometer*" or " Pedometer*" or " inclinometer*" or "Actigraphy" or "activity monitor*" or "step counter").ti,ab,kw. 48048
17 14 or 15 or 16 223804
18 rehabilitation medicine/ or rehabilitation/ or *physiotherapy/ or rehabilitation center/ or *subacute care/ or *postoperative period/ or *aftercare/ or *postoperative care/ 175732
19 (rehab* or e rehab* or Physical Therapy Modaliti* or "physical therap*" or physiotherap* or Rehabilitation Center* or Subacute Care or postacute or post-acute or Postoperative Period or Postoperative Care or post-operative or inpatient or sub-acute or hospital care or aftercare or followup care or follow-up care).ti,ab,kw. 791237
20 18 or 19 844772
21 exp mobile application/ or exp telehealth/ or exp telerehabilitation/ or exp accelerometer/ or exp accelerometry/ or exp pedometer/ or exp actimetry/ or exp activity tracker/ 146547
22 15 or 16 or 21 232224
23 8 and 13 and 20 and 22 1538

**Emcare**

Ovid Emcare <1995 to 2023 Week 20>

1 exp sedentary lifestyle/ 5267
2 exp health behavior/ 132875
3 television viewing/ 1643
4 *screen time/ 487
5 *exercise/ 45538
6 *physical activity/ 35417
7 ("sedentary behavio?r*" or "Sedentary Lifestyle" or "Physical Inactivit*" or "Lack of Physical Activit*" or "Sedentary Time*" or "Sitting" or "screen time*" or "inactive" or "Sedentary" or "watching TV" or "TV watching" or "viewing TV" or "TV viewing" or "television watching" or "watching television" or "television viewing" or "viewing television" or "screen-based " or "media time" or "Physical activit*" or "Exercise").ti,ab,kw. 247503
8 1 or 2 or 3 or 4 or 5 or 6 or 7 373451
9 geriatrics/ 9100
10 exp aged/ 631995
11 exp very elderly/ 28307
12 ("Aged" or "Aged, 80 and over" or "frail elderly" or "elder*" or "aging" or "ageing" or "oldest old" or "older person*" or "old person*" or "older patient*" or "old patient*" or "older women" or "old women" or "older men" or "old men" or "old adult*" or "older adult*" or "Older individual*" or "old people" or "older people" or "septuagenarian*" or "octagenarian*" or "octogenarian*" or "nonagenarian*" or "centenarian*" or "senescence" or "senescent" or "geriatric" or "geriatrics" or "geriatrics" or "senior" or "seniors" or "older population" or "neurogeriatr*" or "Sixty plus" or "Middle Aged" or "60 years old" or "60 age" or "63 years old").ti,ab,kw.639266
13 9 or 10 or 11 or 12 1055411
14 mobile application/ 4577
15 exp telehealth/ 25898
16 exp telerehabilitation/ 747
17 *accelerometer/ 2397
18 *accelerometry/ 1293
19 *pedometer/ 574
20 *actimetry/ 582
21 *activity tracker/ 168
22 ("Appstore" or "App store" or "Google play*" or "Googleplay" or "Playstore" or "Play Store" or "ehealth" or "e-health" or "mhealth" or "m-health" or "e-rehab*" or "e-exercis*" or "personal mobility device*" or (("electronic" or "online" or web or internet) adj (platform* or wearable*)) or "webbased" or "web based" or "digital software" or smartwatch* or ((application* or App or apps) adj (mobile or "smartphone" or smartphone* or Apple or Samsung or Android))).ti,ab,kw. 38044
23 ("Accelerometry" or "Accelerometer*" or " Pedometer*" or " inclinometer*" or "Actigraphy" or "activity monitor*" or "step counter").ti,ab,kw. 19627
24 14 or 15 or 16 or 17 or 18 or 19 or 20 or 21 or 22 or 23 81348
25 rehabilitation medicine/ or rehabilitation/ or *physiotherapy/ or rehabilitation center/ or *subacute care/ or *postoperative period/ or *aftercare/ or *postoperative care/ 77988
26 (rehab* or e rehab* or Physical Therapy Modaliti* or "physical therap*" or physiotherap* or Rehabilitation Center* or Subacute Care or postacute or post-acute or Postoperative Period or Postoperative Care or post-operative or inpatient or sub-acute or hospital care or aftercare or followup care or follow-up care).ti,ab,kw. 263936
27 25 or 26 272396
28 8 and 13 and 24 and 27 400

**Web of Science**
*Search carried out on 30-05-2023*

5
#1 AND #2 AND #3 AND #4
1,226
4
TS=((rehab* or e rehab* or Physical Therapy Modaliti* or "physical therap*" or physiotherap* or Rehabilitation Center* or Subacute Care or postacute or post-acute or Postoperative Period or Postoperative Care or post-operative or inpatient or sub-acute or hospital care or aftercare or followup care or follow-up care))
1,106,185
3
TS=( ("Aged" OR "Aged, 80 and over" OR "frail elderly" OR "elder*" OR "aging" OR "ageing" OR "oldest old" OR "older person*" OR "old person*" OR "older patient*" OR "old patient*" OR "older women" OR "old women" OR "older men" OR "old men" OR "old adult*" OR "older adult*" OR "Older individual*" OR "old people" OR "older people" OR "septuagenarian*" OR "octagenarian*" OR "octogenarian*" OR "nonagenarian*" OR "centenarian*" OR "senescence" OR "senescent" OR "geriatric" OR "geriatrics" OR "geriatrics" OR "senior" OR "seniors" OR "older population" OR "neurogeriatr*" OR "Sixty plus" OR "Middle Aged"[Mesh] OR "60 years old" OR "60 age" OR "63 years old"))
1,767,341
2
TS=("sedentary behavior*" OR "sedentary behaviour*" OR "Sedentary Lifestyle" OR "Physical Inactivit*" OR "Lack of Physical Activit*" OR "Sedentary Time*" OR "Sitting" OR "screen time*" OR "inactive" OR "Sedentary" OR "watching TV" OR "TV watching" OR "viewing TV" OR "TV viewing" OR "television watching" OR "watching television" OR "television viewing" OR "viewing television" OR "screen-based " OR "media time" OR "Physical activit*" OR "Exercise*")
871,949
1
TS=("Appstore" or "App store" or "Google play*" or "Googleplay" or "Playstore" or "Play Store" or "ehealth" or "e-health" or "mhealth" or "m-health" or "e-rehab*" or "e-exercis*" or "personal mobility device*" or (("electronic" or "online" or web or internet) NEAR/3 platform* or wearable*) or "webbased" or "web based" or "digital software" or smartwatch* or ((application* or App or apps) NEAR/3 mobile or "smartphone" or smartphone* or Apple or Samsung or Android) OR "Accelerometry" or "Accelerometer*" or " Pedometer*" or " inclinometer*" or "Actigraphy" or "activity monitor*" or "step counter" OR telerehabilitation OR telehealth OR Actimetry activity tracker*)
361,362

**The Cochrane library**

Search Name:
Date Run: 30/05/2023 20:23:18
Comment:

ID Search Hits
#1 MeSH descriptor: [Rehabilitation] this term only 1341
#2 MeSH descriptor: [Physical Therapy Modalities] this term only 4713
#3 MeSH descriptor: [Rehabilitation Centers] this term only 346
#4 MeSH descriptor: [Subacute Care] explode all trees 35
#5 MeSH descriptor: [Postoperative Period] this term only 6225
#6 MeSH descriptor: [Inpatients] this term only 1383
#7 MeSH descriptor: [Aftercare] this term only 1022
#8 (((rehab* or e-rehab* or Physical Therapy Modaliti* or "physical therap*" or physiotherap* or Rehabilitation Center* or Subacute Care or postacute or post-acute or Postoperative Period or Postoperative Care or post-operative or inpatient or sub-acute or hospital care or aftercare or followup care or follow-up care))):ti,ab 366671
#9 #1 OR #2 OR #3 OR #4 OR #5 OR #6 OR #7 OR #8 372494
#10 MeSH descriptor: [Mobile Applications] this term only 1550
#11 MeSH descriptor: [Telemedicine] this term only 3532
#12 MeSH descriptor: [Accelerometry] this term only 694
#13 MeSH descriptor: [Smartphone] this term only 1008
#14 Appstore OR App store OR Google play* OR Googleplay OR Playstore OR Play Store OR ehealth OR e-health OR mhealth OR m-health OR e-rehab* OR e-exercis* OR personal mobility device* OR ((electronic OR online OR web OR internet) AND (platform* OR wearable*)) OR webbased OR web based OR digital software OR smartwatch* OR (application* OR App OR apps) AND (mobile OR smartphone* OR Apple OR Samsung OR Android) OR Accelerometry OR Accelerometer* OR Pedometer* OR inclinometer* OR Actigraphy OR activity monitor OR step counter:ti,ab 121045
#15 MeSH descriptor: [Sedentary Behavior] this term only 1579
#16 MeSH descriptor: [Health Behavior] this term only 5244
#17 MeSH descriptor: [Exercise] this term only 25434
#18 MeSH descriptor: [Screen Time] explode all trees 67
#19 sedentary behaviour* OR Sedentary Behavior* OR Sedentary Lifestyle OR Physical Inactivity OR Lack of Physical Activity OR Sedentary Time OR Sedentary Times OR Sitting OR screen times OR inactive OR Sedentary OR watching TV OR TV watching OR viewing TV OR TV viewing OR television watching OR watching television OR television viewing OR viewing television OR screen-based OR media time OR Physical activit* OR Exercise:ti,ab 169225
#20 MeSH descriptor: [Frail Elderly] explode all trees 1006
#21 MeSH descriptor: [Aged] explode all trees 255129
#22 MeSH descriptor: [Aged, 80 and over] this term only 62305
#23 MeSH descriptor: [Geriatrics] this term only 398
#24 (elderly OR elder OR elders OR aged OR aging OR ageing OR oldest old OR older person* OR old person* OR older patient* OR old patient* OR older women OR old women OR older men OR old men OR old adult* OR older adult* OR Older individual* OR old people OR older people OR septuagenarian* OR octagenarian* OR octogenarian* OR nonagenarian* OR centenarian* OR senescence OR senescent OR geriatric OR geriatrics OR senior OR seniors OR older population OR neurogeriatr* OR Sixty plus OR 60 years old OR 60 age OR 63 years old OR 63 ±):ti,ab 378945
#25 #20 OR #21 OR #22 OR #23 #24 255220
#26 #15 OR #16 OR #17 OR #18 OR #19 174451
#27 #10 OR #11 OR #12 OR #13 OR #14 123197

| | | |
|---|---|---|
| #28 | #9 AND #25 AND #26 AND #27 | 1154 |

Trials: 818
